# Trends in suicide rates during the COVID-19 pandemic restrictions in a major German city

**DOI:** 10.1101/2020.10.21.20187419

**Authors:** Daniel Radeloff, Rainer Papsdorf, Kirsten Uhlig, Andreas Vasilache, Karen Putnam, Kai von Klitzing

## Abstract

**Background:** It remains unclear whether the COVID-19 pandemic is having an impact on suicide rates. Social distancing, economic insecurity and increasing prevalence rates of mental disorders may cause an increase in risk factors for suicide.

**Methods:** Data on suicide events in a major city in Germany, and the corresponding life years were provided by the local authorities. For the year 2020, periods without restrictions on freedom of movement and social contact were compared with periods of moderate and severe COVID-19 restrictions. To avoid distortions due to seasonal fluctuations in suicide risk, data from 2020 were compared to data from 2010 to 2019.

**Outcomes:** A total of 333 suicides were registered and 2,791,131 life years (LY) were spent between 2010 and 2020. Of these, 42 suicides and 300,834 LY accounted for the year 2020.

In 2020, suicide rates (SR, suicides per 100,000 LY) were lower in periods with moderate (SR = 8.5, χ^2^ = 4.374, p = 0.037) or severe COVID-19 restrictions (SR = 7.0, χ^2^ = 3.999, p = 0.046) compared with periods without restrictions (SR = 18.0). A comparison with preceding years showed that differences cannot be attributed to seasonal variations. No age- or gender differences were found.

**Interpretation:** SR decreased during the COVID-19 restrictions; we expect SR to rise in the medium term. Careful monitoring of SR in the further course of the COVID-19 crisis is therefore urgently needed. The findings have regional reference and should not be over-generalized.

**Funding:** This study was conducted without external funding.

## Introduction

It remains unclear whether the COVID-19 pandemic is having an impact on suicide rates. Some predict that suicide rates will rise, since actions to contain COVID-19, such as social distancing, economic lockdown, or the temporary restructuring of the health system, could cause risk factors for suicide to increase ^1–5^. Indeed, analyses of previous economic crises have shown that an increase in unemployment was associated with an increase in suicide rates ^6–10^; and according to leading theories of suicide prevention, the loss of social inclusion is a major risk factor for suicide ^11^.

In Germany, a significant restriction of the free movement of persons was agreed upon in March 2020, with the strongest restrictions coming into force in April and May. German borders were virtually closed for travel from 16 March onwards ^12–14^. On 22 March 2020, the German Federal Government and the Länder agreed on a comprehensive restriction of social contacts, which required people to reduce contacts with others (except for members of one ‘s own household) to an absolute minimum ^15^. In the Free State of Saxony, further restrictions on going out, and a ban on visiting care homes, were adopted on 1 April 2020 ^16^. While the restrictions on going out were eased on 4 May, the restrictions on visiting care homes remained ^17^. Following a meeting by the EU interior ministers on 15 June, extensive freedom of movement within the EU ‘s Schengen Area was gradually restored, but differentiated travel warnings and quarantine regulations following travel remained in place for parts of it. The German Federal Foreign Office also maintained the existing travel warnings for 160 countries until 31 August 2020 ^18^.

Elderly people, particularly those with severe or multiple underlying health conditions, are threatened by high fatality rates of COVID-19 ^19^, and are therefore particularly affected by the pandemic. Social distancing was practised more consistently by this age group, and older people are more often dependent on help from third parties. Thwarted belongingness and perceived burdensomeness are in turn central risk factors for suicide, according to the interpersonal theory of suicide ^11^. In the case of senior citizens, these risk factors affect an already vulnerable age group with the highest age-related suicide rates ^20^.

This study investigated the influence of social distancing during the COVID-19 restrictions period on suicide rates (SR). We addressed the following hypotheses:

A.SR increased in the total population under conditions compared to periods without restrictions. This applies to both the period of travel restrictions and that of restricting social contact.

B.Within the 70+ age group, the risk of suicide increased more strongly than in other age groups during COVID-19 restrictions.

C.A comparison with previous years shows that differences in SR within 2020 are not due to seasonal differences.

## Methods

### Sample and data acquisition

The data on suicides are based on the City of Leipzig ‘s cause of death statistics, and were provided by the responsible health authority for the years 2010 to 2020. Data were obtained for age ranges 0–4, 5–9, 10–14, …, 75-79, 80–84, 85+ and for both sexes. Annual population statistics were provided for the corresponding age groups by the residents ‘registration office of Leipzig (https://statistik.leipzig.de/statcity/).

### Analytical strategy

Data were analysed using the R software version 3.3.1 ^17^, IBM SPSS 25.0 ^18^, and Microsoft Excel.

The analysis included suicides from the first six months of each of the years studied. For the year 2020, months without restrictions on freedom of movement or social contact were aggregated as period nR_2020 (January, February), those with moderate restriction as period R1_2020 (travel restrictions; March to June), and those with severe restrictions as period R2_2020 (restrictions on travel, going out and social contact; April, May). To compare suicide mortality in 2020 before and during the COVID-19 restrictions, suicide cases were assigned to group nR_2020, R1_2020 and R2_2020. Corresponding life-years were calculated, according to the length of the periods examined.

Life years (LY) and events were used to calculate the Risk Ratios (RR) with Incidence Rate Ratios (IRR). Differences in suicide risk between nR_2020 and the risk groups R1_2020 and R2_2020 were conducted using Chi-Square tests. In order to exclude biases due to seasonal fluctuations in the suicide risk, Mantel-Haenszel statistics (MH, Test for Heterogeneity) were performed to examine the risk of suicide within the 2020 restriction periods and with the paired periods for years 2010-2019. The Test for Heterogeneity examines whether the IRR of n 2×2 tables differ. Post-hoc, Chi-Square tests were used to examine differences for each pair (nR_2020 vs nR_2010/19; R1_2020 vs R1_2010/19; R2_2020 vs R2_2010/19).

In order to examine age-related and gender-related differences within the 2020 restriction periods, suicide risk in nR_2020, R1_2020 and R2_2020 were compared between senior age (70+) vs age group 0–69 and between genders using the MH.

### Ethical considerations

The study was approved by the ethics committee of the medical faculty of the University Hospital Leipzig, Germany (study ID: 272/20-ek) and conducted according to the Declaration of Helsinki. This epidemiological cohort study is based on the death statistics. For methodological reasons, no informed consent can be obtained.

## Results

A total of 2,791,131 LY were spent and 333 suicides were registered during the periods studied. In 2020, 18 suicides (LY: 100,278) were attributed to nR_2020, 17 (LY: 200,556) to R1_2020, and 7 (LY: 100,278) to R2_2020. In the previous years 2010 to 2019, 90 suicides (LY: 930,377) were attributed to nR_2010/19, 208 (LY: 1,860,754) to R1_2010/19, and 91 (LY: 930,377) to R2_2010/19. The suicide rates within the individual periods were 18.0, 8.5, 7.0 in nR_2020, R1_2020, and R2_2020, respectively.

The suicide risk in 2020 was found to be different between nR_2020 and the periods studied of R1_2020 χ^2^ [1; N = 300,869] = 4.374, p = 0.037), and R2_2020 R2 (χ^2^ [1; N = 200,581] = 3.999, p = 0.046).

The difference for nR compared to R1 and R2 also remained in a comparison with the previous years 2010 to 2019 (Test for Heterogeneity; periods nR vs R1: Q [df = 1] = 12.233, p < 0.001; periods nR vs R2: Q [df = 1] = 13.974, p < 0.001). The post hoc analysis showed that the described difference was due to high suicide rates in nR_2020 compared to nR_2010/19. The periods R1_2020 and R2_2020 were not different compared to the previous years.

For details, see Figure 1.

**Figure 1:**
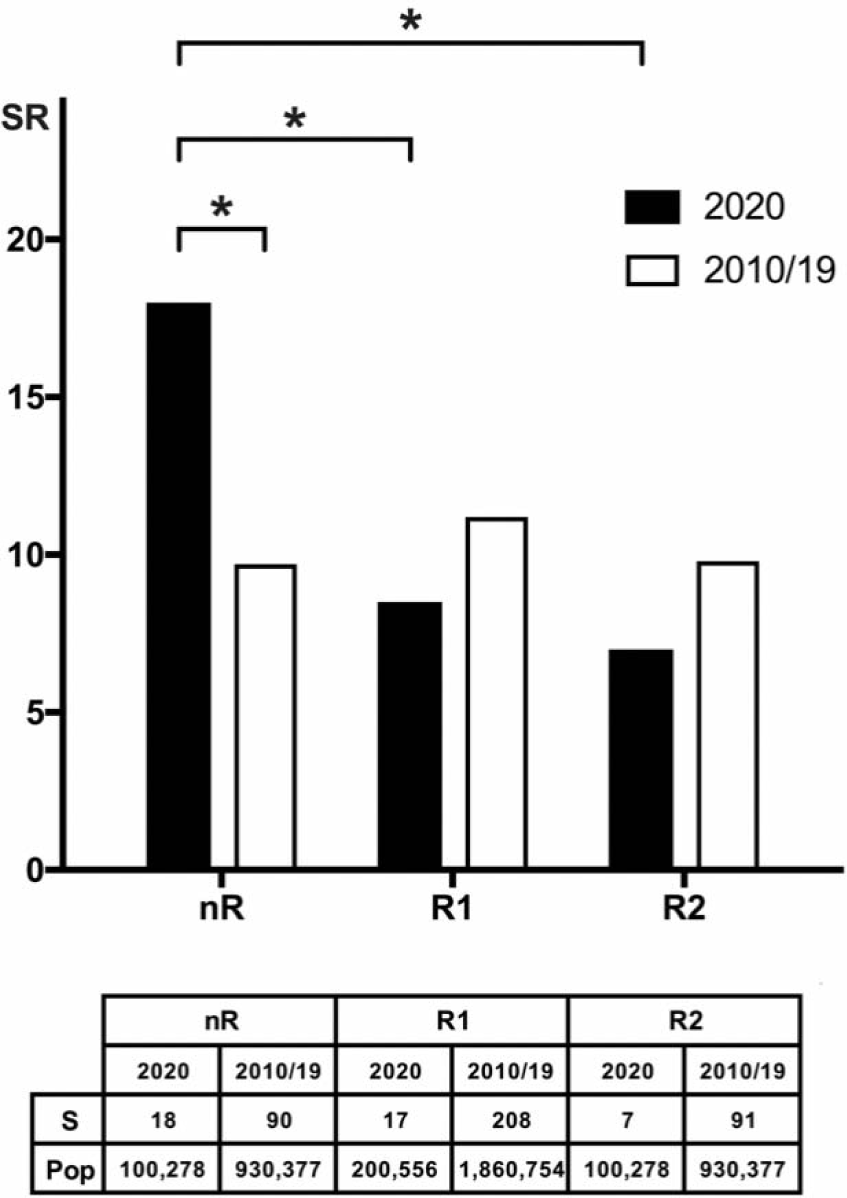
Suicide rates before and during COVID-19 restrictions. This displays the suicide rates (SR, suicides per 100,000 life years) in 2020 (black columns) and the average SR of the years 2010–2019 (white columns). nR = period in 2020 without COVID-19 restrictions (and corresponding periods in previous years), R1 = period of moderate restrictions, R2 = period of severe restrictions. Asterisks indicate significant differences (p < 0.05) in the underlying suicide numbers and population sizes based on Chi-Square statistics. S = suicide numbers, Pop = population sizes (life years).

No gender- or age-related differences were demonstrated between R_2020, R1_2020, and R2_2020.

## Discussion

Our hypotheses regarding suicide rates increasing during corona restrictions could not be confirmed. On the contrary, we found lower suicide rates during the pandemic restrictions in 2020 compared to previous months. However, a comparison with preceding years showed that this difference in 2020 was mainly caused by high suicide rates in the period without restrictions.

Results represent local suicide trends during the COVID-19 pandemic in a major city in Saxony. In Saxony, as in most regions of Germany, the prevalence and mortality rates of COVID-19 were comparatively low, with 135.8 and 251.0 cases per 100,000 inhabitants, respectively ^24^. The regional shut-down was much less restrictive than in other European countries, i.e. United Kingdom, France, Spain or Italy. At an early stage of the pandemic, the German government committed to support measures to prevent insolvencies and unemployment. Accordingly, results should be evaluated under these external conditions. Regional differences in the COVID-19 pandemic may produce regional differences in the mental health situation, economic crises and suicide rates. The findings of this study should not, therefore, be extrapolated uncritically to other regions or countries. It should not be assumed either that the trend described will remain stable. This study only provides a first regional snapshot.

The COVID-19 pandemic is a new phenomenon. Thus, it is not surprising, that little scientific evidence of suicide trends during the COVID-19 pandemic exist. A number of case reports describes a co-occurrence of suicidal behaviour and COVID-19, with the underlying factors being fear of infection ^25–27^ or severe mental disorders such as hallucinations or delusions ^28–30^. However, trends in suicide risks cannot be concluded from case reports or case series.

Projections based on underlying risk factors for suicide, such as unemployment, indicate rising suicide rates during the COVID-19 pandemic ^1,2^. This does not contradict the reduced suicide risk found in our study, since unemployment has not yet risen in the region investigated. It illustrates, however, that suicide rates will not increase immediately, but possibly only with a time lag.

This is consistent with studies reporting no increase in suicidal behaviour, used as an indirect measure for suicide risk, in the early stages of the COVID-19 pandemic ^31–37^. For instance, online surveys showed a decrease in suicidal thoughts and intention during the pandemic; presentation at emergency departments due to suicidal ideation decreased ^36^, suicides in selective autopsy samples remained low ^37^, and search engine users entered suicide-related terms less frequently ^31–33,35^. However, the overall results are inhomogeneous, since other surveys indicate a high prevalence of suicidal thoughts during the pandemic, in particular under quarantine conditions ^3,38–^ 41.

The findings have led us to change our initial hypothesis. We predict the pandemic will see the following course of suicide rates; this will need to be verified in future studies: In an initial phase, the pandemic was perceived as an incalculable new threat without any social countermeasures being taken. In this phase of individual disorientation, suicide rates were unusually high.

In a second phase, with the COVID-19 restrictions, a social response to the threat was given. Rules were established to protect the most vulnerable individuals and to slow down the spread of the virus. The omnipresent concern for the well-being of fellow human beings led to an increased sense of social belonging despite physical distance. According to Durkheim ‘s theory, a temporary increase of social integration and cohesion results in a reduction in suicide rates ^42^. Some findings observed during the Second World War and the 9/11 terror attacks support this hypothesis, but it remains controversial ^43–45^.

In a third phase, the pandemic could be perceived as more predictable and less threatening. The feeling of belonging may once again be increasingly determined by the quality and quantity of everyday contacts, which could continue to be reduced depending on the regulations. A rise in loneliness, unemployment and mental disorders may lead to a delayed increase in the suicide rates in the medium term. As other authors have stressed, it is important to distinguish between physical distance and social belonging ^46^. A major challenge for suicide prevention during the COVID-19 pandemic is to find an answer to the following question: How can we succeed in maintaining a sense of social belonging in our society if physical distance persists in the medium term?

## Conclusion

In the population studied, suicide rates decreased during the COVID-19 restrictions, but we expect suicide rates to increase with a time delay. Careful monitoring of suicide rates in the further course of the COVID-19 crisis is therefore essential. Accordingly, further investigations and meta-analytical approaches are urgently required to monitor suicide rates. In this sense, the available results represent a first step in this direction.

### Limitations and strengths

This study reports first data on suicide trends during the COVID-19 pandemic. The investigated quarantine period covers only six months and the population studied is relatively small with 0.5 M persons. These findings allow conclusions to be drawn for the region and time period investigated. The results do not allow a supra-regional evaluation or assessment of medium-term trends in suicide rates.

## Data Availability

Aggregate data is available on request.

## Acknowledgements

We would like to thank the Leipzig Health Authority for their support and fast provision of relevant data sets. We would also like to thank Monica Buckland for her valuable support in proofreading.

